# Sex Moderates Family History of Alcohol Use Disorder and Childhood Maltreatment Effects on an fMRI Stop-Signal Task

**DOI:** 10.1101/2022.10.31.22281672

**Authors:** Amanda Elton, J. Hunter Allen, Mya Yorke, Farhan Khan, Peng Xu, Charlotte A. Boettiger

## Abstract

**Background:** Childhood maltreatment (CM) and a family history (FH) of alcohol use disorder (AUD) are each associated with increased impulsivity. However, their unique or shared brain targets remain unknown. Furthermore, both CM and FH demonstrate sex-dependent effects on brain and behavior. We hypothesized that CM and FH interact in brain regions involved in impulsivity with sex-dependent effects.

**Methods:** 144 first-year college students (18-19 years old) with varying experiences of CM and/or FH but without current AUD performed an fMRI stop-signal task. We tested interactions between FH, CM, and sex on task performance and blood oxygen level-dependent (BOLD) signal during successful inhibitions. We examined correlations between BOLD response and psychiatric symptoms.

**Results:** Significant three-way interactions of FH, CM, and sex were detected for brain and behavioral data, largely driven by male subjects. In males, CM was associated with poorer response inhibition but only for those with less FH; males with higher levels of both CM and FH demonstrated better response inhibition. Three-way interaction effects on voxel-wise BOLD response during response inhibition were found in bilateral middle frontal gyrus, left inferior frontal gyrus, dorsomedial prefrontal cortex, and posterior cingulate cortex. Network-level analyses implicated the left frontoparietal network, executive control network, and default-mode network. Greater BOLD response in these networks correlated with lower depressive, impulsive, and attentional symptoms, reduced alcohol misuse, greater resilience scores, and heightened trait anxiety.

**Conclusions:** The results highlight sex-divergent effects of heritable and environmental risk factors that may account for sex-dependent expression of psychopathology in response to risk factors.

## INTRODUCTION

Alcohol use disorder (AUD) affects approximately 10% of young adults^1^ and is characterized by a loss of control over the use of alcohol. Risk for AUD is related to both environmental and heritable factors. For example, early life adversity is associated with a dose-related increased risk for alcohol misuse and addiction^2-6^. On the other hand, approximately half of the risk for AUD is heritable^7,8^, and a family history of addiction is especially associated with risk for AUD^5,9^, independent of the adversity related to parental AUD^5^. Substantial evidence further suggests that early life adversity and heritable risk interact to confer vulnerability for AUD. For example, synergistic effects of early life adversity and FH have been found in large samples, indicating that these risk factors produce supra-additive risk for AUD^9,10^. Similarly, synergistic effects of early life adversity and certain genetic polymorphisms on addiction risk have also been demonstrated^11-21^. However, despite the clear relationships between both early life adversity, heritable risk factors, and their interaction on AUD, the neural and behavioral mechanisms by which these two forms of risk contribute to the development of addiction are incompletely determined.

One mechanism by which risk factors may confer vulnerability for AUD is through increased impulsivity. The clinical criteria for AUD strongly implicate impulsivity as an underlying behavioral mechanism. In fact, individuals with AUD demonstrate increased impulsivity on tasks that measure response inhibition, such as stop-signal tasks^22,23^. Furthermore, impulsive behavior in childhood and adolescence, prior to significant substance use, also predicts later life alcohol use^24-26^, suggesting these behaviors precede the development of AUD and represent a neurobehavioral mechanism of risk.

Indeed, individuals with risk factors for AUD demonstrate poorer performance on laboratory tasks of impulsivity compared with low-risk individuals. For example, children and young adults at risk for AUD based on FH demonstrate increased impulsivity on a response inhibition task compared with individuals without this risk factor^27,28^. Adolescents with FH also exhibit altered trajectories of response inhibition neurocircuitry, including middle cingulate, caudate, and middle frontal gyrus, indicating brain mechanisms through which FH might promote risk for AUD^29^. A history of CM also associates with more impulsive responding and functional alterations in particular brain regions. For example, adolescents exposed to childhood neglect show deficits in response inhibition related to increased neural activity in the anterior cingulate cortex, inferior frontal cortex, posterior insula, and striatum relative to low-risk controls^30^. Furthermore, individuals exposed to sexual assaultive trauma in childhood have atypical electrophysiologic developmental trajectories of no-go frontal theta during response inhibition, which associates with increased risk for AUD symptoms^31^. Additionally, during a stop-signal task, adults with a history of abuse and neglect display altered functional connectivity within an inhibitory control network involving the anterior cingulate cortex and inferior frontal cortex^32^. These studies provide emerging evidence for an association between environmental and heritable AUD risk factors and deficits in response inhibition related to alterations in prefrontal circuit function.

Biological sex also significantly affects the relationship between AUD risk factors, impulsivity, and AUD. Not only are there sex-related variations in the healthy brain^33^, including functional differences during response inhibition^34^, but there are also sex differences in the neurodevelopmental effects of traumatic experiences (the rates of which differ between the sexes^35^) on the neural substrates of response inhibition^32^. For example, childhood maltreatment-related alterations in the functional connectivity strength and organization of a neural network implicated in inhibitory control are sexually dimorphic, such that more left (versus right)-lateralized network connectivity impaired response inhibition in males but improved response inhibition in females^32^. Genetic risk for AUD may also vary by sex, perhaps due to sex differences in etiological pathways or patterns of transmission^36^. Furthermore, studies of healthy adults with FH demonstrate significant sex-by-risk interactions on both brain^37^ and behavioral^38^ measures of response inhibition. Differential effects of risk factors in males and females could partly account for sex differences in the expression of AUD and other psychiatric disorders.

Despite mounting evidence that heritable and environmental factors each influence response inhibition behavior and associated neurocircuitry, neuroimaging studies have yet to examine their unique and combined influence on the brain within a single sample. Investigating both factors in a single study is especially important to disentangle their effects since FH, a proxy for heritable risk, is often associated with CM^5^. Furthermore, we sought to examine how effects of risk factors differ by sex and potentially relate to sex differences in the expression of psychiatric symptoms. We hypothesized that greater levels of each risk factor would be associated with reduced inhibitory control, sex-specific patterns of diminished activation in prefrontal brain regions, and their combination would result in supra-additive impacts on brain and behavioral markers of impulsivity. We tested these hypotheses in a sample of 144 male and female first-year college students with varying levels of heritable and environmental risk for AUD. Subjects performed a stop-signal task while undergoing fMRI. Although increased risk was associated with reduced inhibitory control and attenuated neural responses in a subset of at-risk subjects, as hypothesized, our data further reveled sex-dependent brain functional adaptations in response to heritable and/or environmental risk.

## METHODS

### Subjects

We recruited 165 first-year college students for a neuroimaging study with longitudinal follow-up surveys. The study was approved by the University of North Carolina (UNC) Office of Human Research Ethics and participants gave written informed consent to participate. All participants were 18-19 years of age and in their first year in a 4-year undergraduate degree program at UNC or other colleges in the surrounding region. However, the majority (n=140) were students at UNC. Potential subjects were excluded for MRI contraindications such as claustrophobia and nonremovable metal in the body, as well as left-handedness, psychoactive medication or other routine drug use, neurological disorders, and psychiatric disorders. Lifetime mood or anxiety disorders without meeting criteria currently were not excluded. The Mini-International Neuropsychiatric Interview (M.I.N.I.) for DSM-IV (Sheehan et al., 1998) assessed for the presence of psychiatric disorders, whereas DSM-5 criteria was used to assess current or lifetime AUD or substance use disorder. No participants tested positive on a urine drug screen (Biotechnostix, Inc., Markham, ON) for recreational substance use (including cocaine, cannabis, opioids, amphetamines, methamphetamine) on the day of the MRI scan. Alcohol breathalyzer tests (FC-10, Lifeloc Inc., Wheat Ridge, CO) were similarly negative. 21 subjects had incomplete fMRI or behavioral stop-signal task data and were excluded from the reported analyses, providing a final sample size of 144 subjects (95 females). Of the 144 subjects included in the current analysis, 120 (82 females) completed one-year follow-up surveys of their recent substance use.

### Self-Report Instruments

Self-report questionnaires were administered via Research Electronic DATA Capture (REDCap)^39^ surveys sent to subjects via email. FH was assessed with the Family History Assessment Module (FHAM)^40^. A FH density score was calculated from the total prevalence of AUD among biological parents and second-degree relatives with AUD previously published methodology^41^. The score is a weighted total of relatives with an AUD, where affected parents are weighted 0.5, grandparents are weighted 0.25, and maternal and paternal aunts and uncles are each weighted 0.25 divided by the total number of aunts and uncles on that side of the family.

Childhood maltreatment was measured with the Childhood Trauma Questionnaire (CTQ)^42^. The CTQ includes subscales pertaining to physical, emotional, and sexual abuse, as well as physical and emotional neglect. A log-transformed total score of all five subscales was used in primary analyses. Secondary analyses explored effects of individual subscales.

Baseline surveys also measured ADHD-related hyperactivity and inattention (Conners Adult ADHD Rating Scale [CDDR]^43^) and psychological resilience (Connor-Davidson Resilience Scale [CD-RISC-10]^44^). Both baseline and one-year follow-up surveys included self-report measures of depression (Beck Depression Inventory [BDI]^45^), anxiety (State-Trait Anxiety Inventory [STAI]^46^), alcohol misuse (Alcohol Use Disorders Identification Test [AUDIT]^47^), and other substance use (Customary Drinking and Drug Use Record [CDDR]^48^).

### fMRI

FMRI data during the stop signal task were acquired with a Siemens 3T Prisma scanner with a 32-channel TEM send-receive radio frequency (RF) head coil (Siemens Healthineers, Erlangen, Germany). Blood oxygenation level-dependent (BOLD) images were acquired in sagittal orientation using a multiband EPI sequence with multiband factor=8, TR=800 ms, TE=37 ms, flip angle=52°, 2 mm isotropic voxels, 72 slices, FOV=208×208, bandwidth=2290 Hz/pixel, interleaved acquisition. Due to a scanner update, the sequence was adjusted for 27 subjects to bandwidth= 2186 Hz/pixel and TE=38.2 ms, which would negligibly affect the BOLD contrast. We used an anterior-to-posterior (AP) phase encoding direction for the first 413 TRs and posterior-to-anterior (PA) phase encoding for final 413 TRs. The second scan started upon the completion of the first scan, providing a continuous scan of approximately 11 min.

High-resolution magnetization-prepared rapid gradient-echo (MPRAGE) T1-weighted parameters were: TR= 2530 ms, TE=2.3 ms, flip angle=9°, 1 mm isotropic voxels, 176 sagittal slices, FOV=256×256.

### Stop Signal Task

The stop signal task was presented with Psychopy software, and consisted of 300 trials, divided into two runs corresponding with the AP and PA phase encoding directions of the fMRI scan. Within each run were three 100 s blocks of 50 trials each flanked by four 12 s blocks of fixation. There was a 2000 ms fixed interval between trials. At the start of each trial, an arrow appeared in the center of the monitor with an equal probability of facing left or right. Subjects were instructed to make a left or right button press corresponding with the arrow direction. In 75 trials (25%), a stop signal, an “X”, appeared overtop the arrow after a brief delay, the stop-signal delay (SSD). Subjects were instructed to attempt to withhold their response if the X appeared. The SSD started at 200 ms and was adaptively lengthened by 50 ms after each successful stop and shortened by 50 ms after a failure to withhold a response. The SSD was restricted to a minimum of 50 ms and a maximum of 900 ms. 25 trials (8%) were “null” trials consisting of a bidirectional arrow visual stimulus to aid in deconvolving BOLD signals. Trial order was pseudorandomized. Speed and accuracy were equally emphasized, and subjects were reassured that it was normal to not be able to inhibit their response on every stop trial. Subjects underwent a 50-trial practice prior to entering the scanner, and subjects were reminded to not to wait for the stop signal.

### Behavioral Analysis

We applied lenient criteria for identification of outlier subjects based on published recommendations (inhibition rate on stop trials outside 25-75%, go rate<60%, go accuracy<90%, SSRT<50 ms^49^), which resulted in the exclusion of one subject with a negative SSRT. Despite explicit instructions, numerous subjects displayed evidence of employing waiting strategies. Because the longest SSD included in the task was 900 ms, we further excluded 20 subjects (from behavioral analyses only) with prolonged response times that averaged greater than 950 ms (maximum SSD + lower limit SSRT).

We tested effects of FH, CM, and sex, as well as their interactions, on SSRT in linear regression models. Model 1 simultaneously tested effects of FH and effects of CM, controlling for sex. Model 2 tested interacting effects of FH and CM, controlling for sex. Model 3 testing interacting effects of FH and sex, as well as interacting effects of CM and sex, within the same model. Model 4 tested the three-way interaction of FH, CM, and sex

### fMRI Preprocessing

Data were preprocessed with fMRIPrep^50^. The T1-weighted (T1w) image was corrected for intensity non-uniformity (INU) with *N4BiasFieldCorrection* in ANTs 2.3.3. The T1w-reference was then skull-stripped with a *Nipype* implementation of *antsBrainExtraction*.*sh* workflow (from ANTs), using OASIS30ANTsas target template. Brain tissue segmentation of cerebrospinal fluid (CSF), white-matter (WM) and gray-matter (GM) was performed *recon-all* (FreeSurfer 6.0.1), and the brain mask estimated using *fast* (FSL 5.0.9). Brain surfaces were reconstructed using *recon-all*, and the brain mask estimated previously was refined with a custom variation of the method to reconcile ANTs-derived and FreeSurfer-derived segmentations of the cortical graymatter of Mindboggle. Volume-based spatial normalization to one standard space (MNI152NLin2009cAsym) was performed through nonlinear registration with *antsRegistration*, using brain-extracted versions of both T1w reference and the T1w template (*i*.*e*. ICBM 152 Non-linear Asymmetrical template version 2009c).

BOLD preprocessing included motion estimation and realignment, slice time correction, distortion correction, registration to the T1 image, normalization to MNI space, and confounds estimation. First, a reference volume and its skull-stripped version were generated using a custom methodology of fMRIPrep. A B0-nonuniformity map was estimated based on two echoplanar imaging (EPI) references with opposing phase-encoding directions, with *3dQwarp* (AFNI). Based on the estimated susceptibility distortion, a corrected EPI reference was calculated for a more accurate co-registration with the anatomical reference. The BOLD reference was then co-registered to the T1w reference using *bbregister* (FreeSurfer) which implements boundary-based registration. Co-registration was configured with six degrees of freedom. Head-motion parameters with respect to the BOLD reference (transformation matrices, and six corresponding rotation and translation parameters) are estimated before any spatiotemporal filtering using *mcflirt* (FSL 5.0.9). BOLD runs were slice-time corrected using *3dTshift* (AFNI). The BOLD time-series were resampled into Montreal Neurological Institute standard space (*i*.*e*., MNI152NLin2009cAsym).

### Voxel-wise fMRI Activation Analysis

Voxel-wise activation analysis was conducted using the general linear modelling (GLM) approach^51^ with 3dDeconvolve and 3dREMLfit in AFNI software (version 20.3.00). We modeled the follow trial types: successful stops, failed stops, go trials following successful stops, go trials following failed stops, go trials not following a stop trial (subsequently referred to as “Go trials”), and missed go trials. Nuisance signals were included as covariates, including 24 motion parameters^52^, signals from WM and CSF voxels as well as their derivatives, squared values, and square of their derivatives, as well as the first 10 aCompCor components from the combined WM and CSF mask^53^. Time points with a framewise displacement of >0.5 mm were censored. We calculated the contrast map of successful stops minus go trials (SS-Go) to isolate inhibition-related BOLD responses for each individual.

Group-level effects of risk factors on the voxel-wise contrast of SS-Go were assessed with 3dMEMA in AFNI. To evaluate the independent and interacting effects of FH, CM, and sex, we tested four separate models, corresponding to Models 1-4 described in the *Behavioral Analysis* section above including three-way interactions of FH, CM, and sex, as well as main effects and lower-order interactions. All analyses also included Go trial response rate as a covariate since this metric is associated with a greater likelihood of successful stop trials that occur as a result of a general decrease in overall response frequency and is likely to influence the estimate of response inhibition-related brain activation. The cluster-size threshold required for multiple comparisons correction was calculated with 3dClustSim in AFNI after estimating the spatial autocorrelation with 3dFWHMx. Because we tested 4 different models, we set α=0.0125 to correct for the increased probability of detecting significant clusters, the p-value threshold was set to 0.005, and the calculated cluster size threshold was 72 contiguous voxels.

### Network fMRI Analyses

To reduce the number of tests performed and to aid in interpreting results, we also performed analyses on large-scale neural networks. Network-level analyses were conducted using masks of 10 BrainMap networks derived from task activation analyses that were well-matched to resting-state networks^54^ (http://www.fmrib.ox.ac.uk/datasets/brainmap+rsns/). All voxels having a z-score value >3.0 were included in each binary mask.

To calculate network activation during response inhibition, we calculated the mean SS-Go contrast value within each network mask for each subject. Given significant effects of Model 4 for behavior (i.e., SSRT) and voxel-wise maps of the SS-Go contrast, our network-level analyses similarly tested the three-way interaction effects of FH, CM, and sex, covarying for go trial response rate, on SS-Go contrast values for each network. P-values were FDR corrected for multiple tests.

Additionally, given previously-reported effects of CTQ scores and sex on functional connectivity during a stop-signal task^32^, we also performed a beta-series functional connectivity analysis^55^ to examine effects of risk factors and sex on functional interactions between large-scale networks. First, we reran the AFNI GLM analyses, modeling each successful stop and each Go trial separately. Beta estimates for each trial for both of these trial types were calculated in a voxel-wise manner across the whole brain, and the mean value was subsequently extracted within each binary network mask. This produced a beta series for successful stops and a beta series for Go trials for each network. We focused our functional connectivity analysis on the three networks that were significant for the network-level task activation analysis described above. We calculated the correlations among these three networks during successful stops and Go trials separately, representing network-level functional connectivity during each trial type. To compare functional connectivity during successful stops with Go trials, a SS-Go contrast was computed as the difference in Fisher-z transformed correlations for SS and Go trials for each pair of networks.

### Relationships between Brain Measures and Psychiatric Symptoms

To test whether variations in task-related network activation associated with risk factors and sex represent sex-dependent negative consequences or positive adaptations, we tested the multivariate relationship between network-level SS-Go contrast estimates and self-reported psychiatric symptoms (see below for details). Specifically, a canonical correlation analysis in SAS (version 9.4) Proc Cancorr tested for multivariate correlations between brain networks and psychiatric symptoms, identifying linear combinations of these two sets of variables (i.e., brain networks and psychiatric symptoms) to maximize their correlation.

For brain network measures, we included SS-Go activation estimates for the networks that displayed significant effects of CM, FH, and sex in the analysis described above (see *Network fMRI Analyses*). For psychiatric symptom measures, we included self-reported symptoms reported at baseline of depression (BDI), trait anxiety (STAI), ADHD-related hyperactivity and inattention (CAARS), and psychological resilience (CD-RISC-10), as well as substance use measures collected at 1-year follow-up, including marijuana use (0=never used, 1=ever used), tobacco use (0=never used, 1=ever used), and alcohol use (AUDIT total score). The analysis covaried for AUDIT scores at baseline (to model change in drinking), FH, CM, sex, whether baseline assessments were conducted during the Covid-19 pandemic, and whether 1-year follow-up assessments were conducted during the Covid-19 pandemic.

## RESULTS

### Demographic and Questionnaire Data

Characteristics of subjects included in the analyses based on self-report questionnaires are presented in Table 1. The mean CTQ total scores in this sample (Table 1) were somewhat higher than reported in a community sample^56^, as expected due to our targeted recruitment. The sexual abuse subscale of the CTQ was higher in females based on a two-sample t-test, although rates were generally low (Table 1). A chi-square test indicated that by the follow-up, females were more likely to have used marijuana than were males. There were no other sex differences in self-reported data.

**Table 1.**
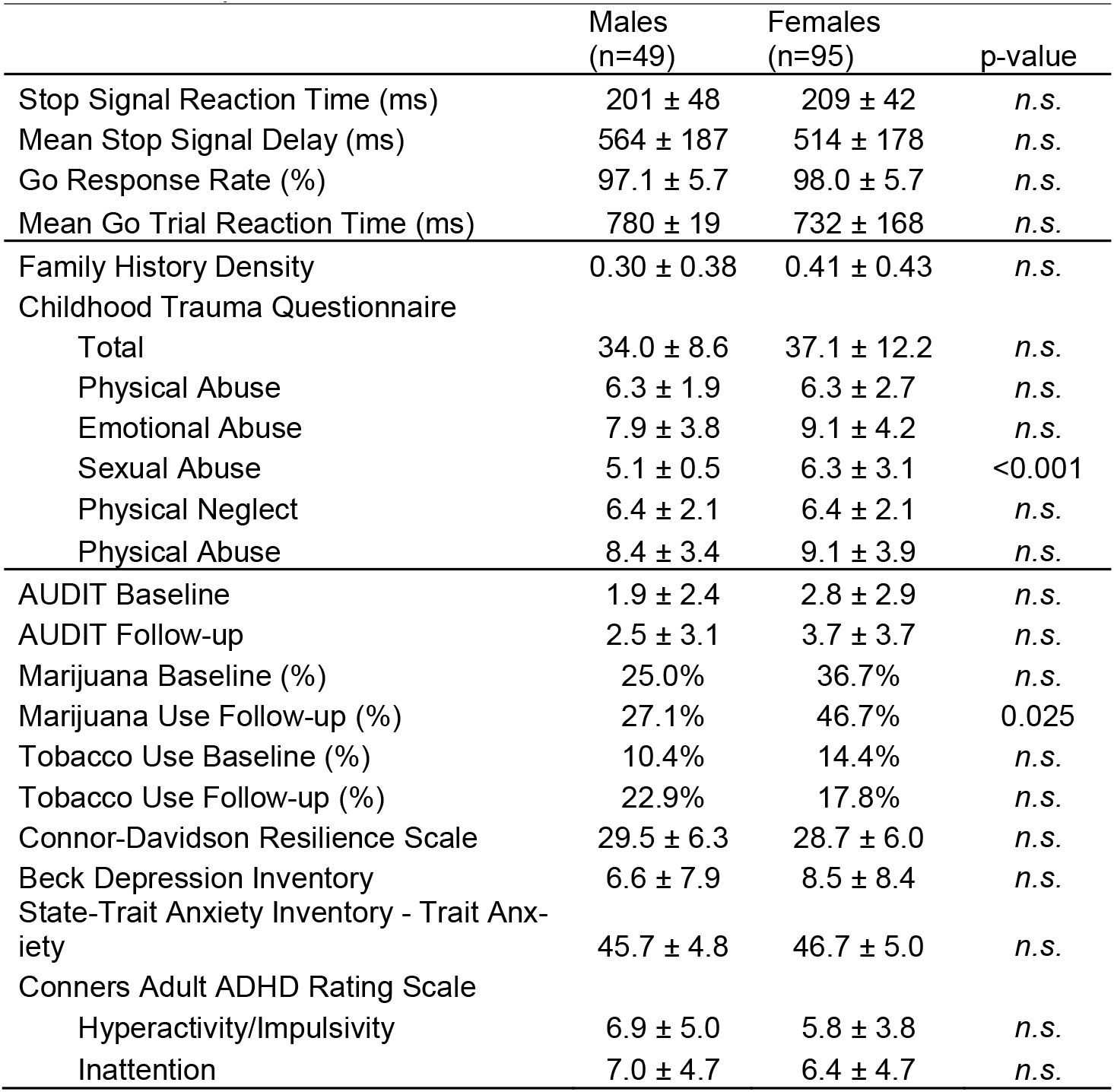
Self-report Questionnaire Data for Males and Females

### Behavioral Data

Stop-signal task performance measures are presented in Table 1. A linear regression identified a significant FH×CM×sex interaction on SSRT (t=2.88, p=0.004). As shown in Figure 1, the plotted data indicated that in men with low levels of FH density, greater CTQ scores are associated with slower SSRTs. On the other hand, greater CTQ scores are associated with faster SSRTs in men with higher FH density. The pattern was opposite in females: Higher CTQ scores and lower FH density predicted faster SSRTs, whereas higher CTQ scores and higher FH density predicted slower SSRTs. Main effects of sex, FH, and CM and lower-order interactions were not significant in models testing these effects.

**Figure 1.**
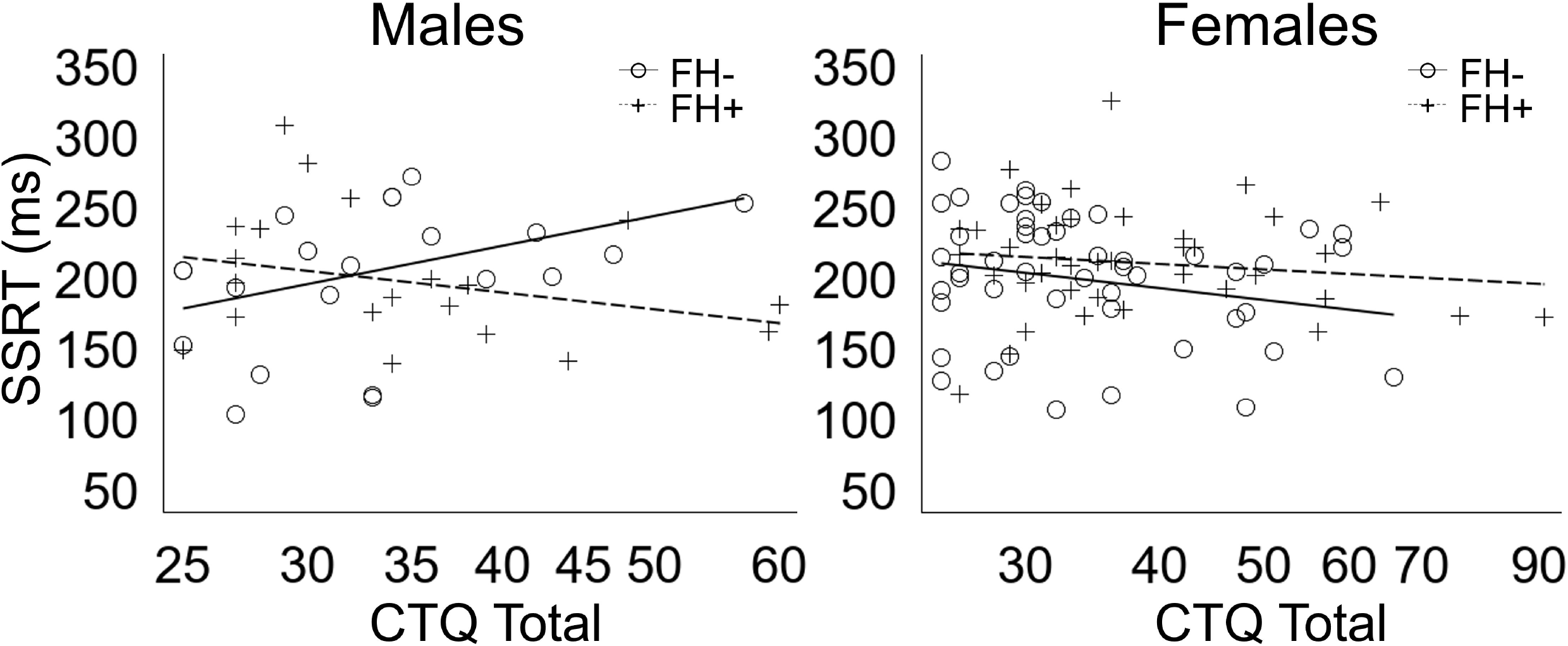
Scatter plots of the significant effects of Childhood Trauma Questionnaire (CTQ) total scores on stop signal reaction times (SSRT) by family history density (FH) and sex. For visualization purposes, FH was binarized such that subjects with a family history density ≤0.25 were designated as FH- and >0.25 was designated as FH+.

### Voxel-wise fMRI Activation Analysis

Results from the voxel-wise analysis of FH, CTQ on the SS-Go contrast are presented in Figure 2 and Table 2. Figure 2a presents regions significantly active in the SS-Go contrast in a 1-sample test of all subjects in the study. Models 1-3 produced no significant clusters. However, Model 4, which tested the three-way interaction of FH, CM, and sex, indicated multiple significant cortical clusters, including in medial prefrontal cortex, bilateral middle frontal gyrus, left inferior frontal gyrus, posterior cingulate cortex, and angular gyrus (Figure 2B). Scatter plots of these findings (Figure 2C) indicate that CTQ scores were associated with lesser task-related activation (i.e., SS-Go contrast) in males with lower FH density but greater task-related network activation in males with higher FH density; in females, CTQ scores were associated with lesser network task-related activation for females with higher FH density.

**Table 2.** Significant Effects of the Interaction of Family History Density, Childhood Maltreatment, and Sex on the Voxel-Wise Contrast of Successful Stops minus Go Trials

| Brain Region | n voxels | x | y | z |
| --- | --- | --- | --- | --- |
| Left Inferior Frontal Gyrus (Pars Orbitalis) | 389 | -42.1 | 27.1 | -19.2 |
| Right Superior Frontal Gyrus | 254 | 12.9 | 55 | 41.7 |
| Left Superior Frontal Gyrus | 254 | -16.6 | 41 | 45 |
| Right Middle Frontal Gyrus | 209 | 41.9 | 20.9 | 52.5 |
| Medial Prefrontal Cortex | 173 | -2.9 | 44.5 | 32 |
| Left Inferior Temporal Gyrus | 129 | -64.4 | -51.4 | -11 |
| Left Middle Frontal Gyrus | 127 | -46.4 | 45.3 | 2.5 |
| Medial Prefrontal Cortex | 122 | -6.4 | 63 | 5.1 |
| Left Angular Gyrus | 113 | -46.9 | -60.5 | 25.9 |
| Posterior Cingulate Cortex | 109 | 6.2 | -40.4 | 34.9 |
| Right Inferior Frontal Gyrus (pars triangularis) | 106 | 56.7 | 34.6 | 13.7 |
| Left Middle Frontal Gyrus | 90 | -40.9 | 10.6 | 56.9 |
| Left Occipital Pole | 88 | -29.2 | -99.9 | -13.4 |
| Right Cerebellum | 84 | 21.6 | -90.1 | -31.4 |
| Right Superior Frontal Gyrus | 80 | 9.4 | 36.7 | 54.7 |
| Left Middle Occipital Gyrus | 74 | -44.6 | -75.7 | 37.4 |

**Figure 2.**
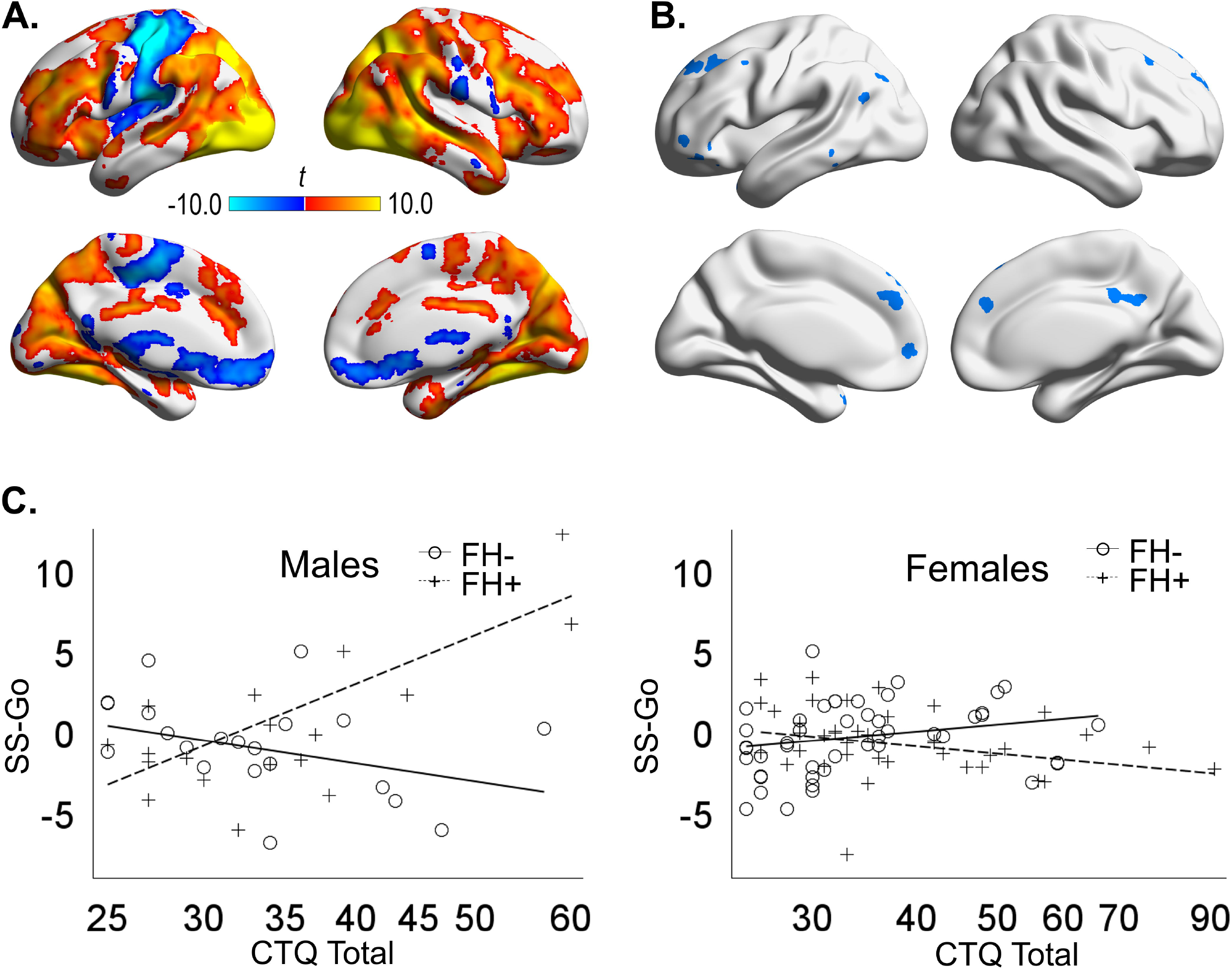
Voxel-wise neuroimaging results. A) Significant clusters from a one-sample t-test of the contrast between successful stop trials and go trials across all subjects. B) Significant clusters of the three-way interaction effect of family history density, Childhood Trauma Questionnaire (CTQ) total scores, and sex on the contrast between successful stop trials and go trials (SS-Go). C) Scatter plots of the effects of CTQ total scores on the SS-Go contrast by family history density (FH) and sex on; plotted values represent the first principal component of all significant clusters. For visualization purposes, FH was binarized such that subjects with a family history density ≤0.25 were designated as FH- and >0.25 was designated as FH+.

### Network fMRI Analyses

Of the ten networks tested, three-way interaction effects of FH, CM, and sex were detected for three networks: the default-mode network (Figure 3A; t=-3.03, p=0.003), executive control network (Figure 3B; t=-2.94, p=0.004), and left frontoparietal network (Figure 3C; t=-2.76, p=0.007). These effects survived FDR correction for multiple tests. The results from these analyses mirrored those in Figure 2B from the whole-brain voxel-wise analysis in which there were sex differences in the effects of CTQ scores on task-related network activation depending on FH density.

**Figure 3.**
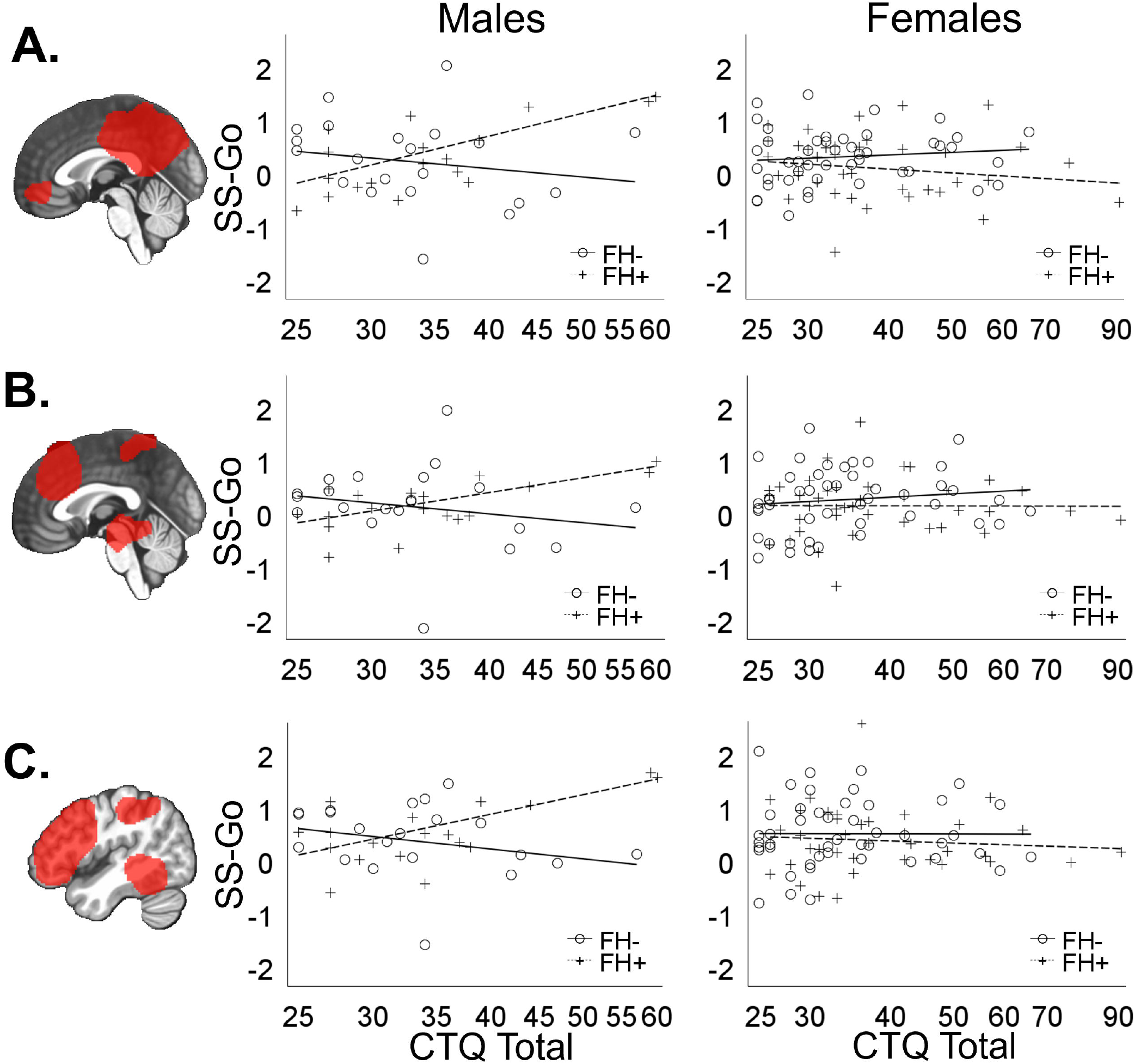
Network-level neuroimaging results. Scatter plots of the effects of Childhood Trauma Questionnaire (CTQ) total scores on the contrast between successful stop trials and go trials (SS-Go) for the A) default-mode network b) executive control network, and C) left frontoparietal network by family history density (FH) and sex. For visualization purposes, FH was binarized such that subjects with a family history density ≤0.25 were designated as FH- and >0.25 was designated as FH+.

We next explored whether effects of risk factors on network activation are confounded by their effects on task performance or substance use. First, we controlled for SSRT by including it as a predictor in the linear regression model testing effects of FH, CM, and sex on network activation. Three-way interaction effects of FH, CM, and sex remained significant for the default-mode network (t=-2.68, p=0.008), executive control network (t=-3.01, p=0.003), and left frontoparietal network (t=-3.41, p<0.001). We also tested these effects when controlling for substance use, represented by AUDIT total score, marijuana use (yes/no), and tobacco use (yes/no). Again, three-way interaction effects remained significant for the default-mode network (t=-3.29, p=0.001), executive control network (t=-2.82, p=0.006), and left frontoparietal network (t=-3.12, p=0.002).

Testing for effects of risk factors and sex on task-related functional connectivity, a beta-series correlation analysis analyzed the functional connectivity during successful stops versus Go trials between the default-mode network, executive control network, and left frontoparietal network. There were no significant main or interaction effects of FH, CM, and sex on task-dependent functional connectivity between these networks, even at an uncorrected p<0.05.

### Brain Network Relationships with Behavioral Symptoms

A canonical correlation analysis examined relationships between SS-Go activation estimates of the networks demonstrating significant interaction effects of FH, CM, and sex (i.e., default-mode network, executive control network, and left frontoparietal network) and multiple psychiatric measures. There were two significant correlations between brain network and psychiatric measures (canonical correlation 1=0.43, p=0.002; canonical correlation 2=0.39, p=0.022). To aid in interpreting the canonical correlations, the correlations between the raw variables for the brain and the canonical variates for the brain as well as correlations between the raw variables for psychiatric measures and the canonical variates for psychiatric measures are presented in Table 3. The first canonical correlation indicates that greater activation of the executive control and left frontoparietal networks correlated with general reductions in psychopathology across ADHD, anxiety, depression, and substance use domains. A second canonical correlation indicated that increased default-mode network activation, coupled with activation of executive control and left frontoparietal networks, correlated with greater psychological resilience and reduced ADHD and depressive symptoms, but also with heightened anxiety.

**Table 3.** Variable Correlations with Canonical Variates for Network Activation and Psychiatric Symptoms

|  |  | *Canonical Correlation 1* | *Canonical Correlation 2* |
| --- | --- | --- | --- |
| *Brain Variables* | Network 4 (DMN) | -0.09 | 0.99 |
|  | Network 8 (ECN) | 0.39 | 0.79 |
|  | Network 10 (LFPN) | 0.70 | 0.71 |
| *Psychiatric Variables* | Audit Score | -0.22 | 0.02 |
|  | Cannabis Use | -0.46 | -0.22 |
|  | Tobacco Use | -0.63 | 0.28 |
|  | Inattentive Symptoms | -0.63 | -0.51 |
|  | Hyperactive-Impulsive Symptoms | -0.43 | -0.48 |
|  | Depression Symptoms | -0.30 | -0.48 |
|  | Trait Anxiety | -0.29 | 0.41 |
|  | Resilience | -0.10 | 0.53 |

Pairwise correlations, covarying for identical variables as the canonical correlations, were also calculated to supplement the canonical correlation analysis. Pearson correlations and uncorrected p-values from theses univariate analyses are reported in Table 4 and largely mirror results of the multivariate correlation approach.

**Table 4.** Pairwise Correlations between Brain Networks and Psychiatric Symptoms

|  | Network 4<br>(DMN) | Network 8<br>(ECN) | Network<br>10 (LFPN) |
| --- | --- | --- | --- |
| **Audit Score** | 0.00<br>*0.99* | -0.04<br>*0.71* | -0.06<br>*0.58* |
| **Cannabis Use** | -0.04<br>*0.71* | -0.06<br>*0.53* | -0.19<br>*0.07* |
| **Tobacco Use** | 0.11<br>*0.30* | -0.09<br>*0.40* | -0.12<br>*0.25* |
| **Inattentive Symptoms** | -0.20<br>*0.06* | **-0.35**<br>*<0.001* | **-0.40**<br>*<0.001* |
| **Hyperactive-Impulsive Symptoms** | -0.18<br>*0.09* | **-0.22**<br>*0.03* | **-0.32**<br>*0.001* |
| **Depression Symptoms** | -0.19<br>*0.07* | **-0.32**<br>*0.002* | **-0.28**<br>*0.01* |
| **Trait Anxiety** | **0.20**<br>*0.05* | 0.07<br>*0.48* | 0.02<br>*0.84* |
| **Resilience** | **0.24**<br>*0.02* | **0.24**<br>*0.02* | 0.16<br>*0.13* |

Pearson correlations and p-values (*italics*) are reported for the pairwise correlation between brain networks and psychiatric symptoms. DMN, default-mode network; ECN, executive control network; lfpn, left frontoparietal network

## DISCUSSION

Although we predicted the interaction of FH and CM would be associated with supra-additive adverse effects on behavioral and neural measures of response inhibition, analyses revealed potential positive adaptations associated with greater risk exposure that were sex-dependent. Effects of risk factors were more pronounced in male subjects, in which CM was associated with heightened impulsivity and reduced BOLD response in males without FH; However, males with higher levels of both risk factors were more likely to exhibit better behavioral performance and greater BOLD response during response inhibition. Effects in females were overall smaller but trended in the opposite direction as males. Additionally, whereas greater engagement of the executive control network and left frontoparietal network during the stop-signal task was associated with reduced symptoms of ADHD and depression, default-mode network engagement was associated with both increased trait anxiety and higher resilience scores. The findings suggest the potential for sex-specific adaptations in response to risk and indicate brain mechanisms by which males and females may differ in the prevalence and expression of different forms of psychopathology.

### Behavioral Analysis

Prior studies have pointed to poor response inhibition in individuals with FH^27,28^ and childhood adversity^30^, although behavioral effects are not consistently found (e.g., ^32,57^). Our results highlight the complexity of effects of risk factors and sex on SSRT. Specifically, the data indicate males with increased levels of both heritable and environmental risk for AUD demonstrate a more “resilient” behavioral phenotype (i.e., preserved SSRT) compared with males with only moderate levels of risk, whereas minimal effects were seen in females (Figure 1). The observed interaction effect mirrors previous findings in at-risk young adult alcohol drinkers who were otherwise healthy, which showed a significant sex-by-FH interaction in which men with familial risk had improved inhibitory control relative to at-risk females^38^. An interpretation of these findings is that high-risk males who do not meet criteria for any psychiatric disorders at 18-19 year of age tend to exhibit positive adaptations such as preserved response inhibition. On the other hand, this behavioral measure may not be as strong of an index of AUD risk or resilience in females.

### Task activation

Analyses detected risk- and sex-dependent increases and decreases in BOLD response across the cortex in regions associated with the left frontoparietal network, executive control network, and default-mode network. Greater BOLD response during response inhibition has been interpreted as reflecting either compensatory adaptations or negative consequences^58^. Previous studies have demonstrated that, prior to significant substance use, at-risk youth with FH exhibit reduced prefrontal activation during response inhibition, particularly in the dorsal anterior cingulate cortex and lateral prefrontal cortex^29,59^. However, there are also data supporting greater activation of the bilateral middle frontal gyrus and rostral anterior cingulate in adolescents with FH during Stroop interference^60^. Reduced BOLD response during response inhibition in adolescence has been associated with future heavy alcohol use^61^, supporting the conclusion that enhanced BOLD response is generally protective. The association of greater activation of the executive control network and left frontoparietal network with lower trajectories of alcohol use (Table 3) further supports this conclusion. Thus, increases or decreases in BOLD response during response inhibition may partly reflect individual differences in vulnerability or resilience.

In the current study, AUD risk was associated with alterations in the left frontoparietal network including the left inferior frontal gyrus. Consistent with the current findings, a previous neuroimaging study in males and females with or without FH reported enhanced recruitment of the left inferior frontal gyrus during response inhibition in at-risk individuals^37^. In both that study and the current study, the effect that was driven by males, with high-risk males demonstrating enhanced recruitment of the left inferior frontal gyrus. The current study adds further evidence that effects on this region are driven by a combination of both heritable and environmental risk factors. Similarly, a study in at-risk adults with histories of childhood maltreatment indicated sex differences in left-versus-right-lateralized functional connectivity during a stop-signal task and corresponding sex differences in the connectivity relationships to SSRT and ADHD symptoms. Response inhibition tends to rely more on the right inferior frontal gyrus^62^, and greater recruitment of the left inferior frontal gyrus has been interpreted as compensatory^37^. Indeed, we observed that males with higher levels of both heritable and environmental risk also exhibited greater activation of the left frontoparietal network and preserved SSRTs, consistent with these brain changes representing risk-related compensatory adaptations.

The executive control network, including two clusters in the medial prefrontal cortex, also demonstrated a significant interaction of FH, CM, and sex. The medial prefrontal/anterior cingulate region represents a major hub of an inhibitory control network that also include the bilateral anterior insula/inferior frontal gyrus^32^. As noted above, a previous study demonstrated sex differences in the effects of childhood maltreatment on the functional connectivity between the anterior cingulate cortex and left and right anterior insula/inferior frontal gyrus, suggesting an important role of this brain region in sex-by-risk factor interactions on inhibitory control. In fact, the anterior cingulate cortex has been implicated in resilience measured with CD-RISC^63, 64^. The current study further suggests a role for this brain region in sex-specific functional adaptations in response to both stress and heritable risk factors.

Our data also indicated increased (males with higher FH and higher CM) or decreased (males with either higher FH or higher CM) activation in regions associated with the default-mode network (Figures 2B, 3), including medial prefrontal cortex and posterior cingulate cortex. Default mode network activity is typically suppressed during externally-oriented tasks to such as the stop-signal task^66^. In fact, in the SS-Go contrast for the whole sample (Figure 2A), default-mode-related regions generally showed non-significant (e.g., posterior cingulate cortex) or negative (ventromedial prefrontal cortex) activation. However, the default-mode network is involved in monitoring internally-represented information^67^. Variations in task-induced BOLD response in these regions related to risk factors and sex may reflect altered vigilance processes during task performance, an inference supported by the positive canonical correlation that highly weighted this network and anxiety symptoms (Table 3). Furthermore, the finding that greater default-network activation was also associated with higher resilience scores is consistent with the possibility that this effect represents a marker of resilience related to AUD risk factors.

Secondary analyses that included SSRT as a covariate indicated that the effects of risk factors and sex on network activation were not driven by the differences in behavioral performance. Furthermore, other analyses suggested that brain effects were not related to subject-level variation in prior substance use. This supports the contention that these brain changes are more closely associated with exposure to risk factors than to the behavioral consequences of those risk factors.

### Brain relationships to Psychiatric Symptoms

Consistent with the notion that lesser activation during response inhibition is associated with greater risk for poor substance use outcomes, a canonical correlation analysis linked lower BOLD response in the executive control and left frontoparietal networks not only with greater trajectories of alcohol use, but also greater incidence of marijuana and tobacco use at follow-up, greater symptoms of inattention and impulsivity, and greater depression (Table 3). However, note that the univariate correlation between networks and change in AUDIT scores was not significant (Table 4). On the other hand, a second canonical correlation indicated that greater engagement of all three networks, especially the default-mode-network, was associated with some positive outcomes such as lower levels of depression and ADHD and higher resilience scores; however, these same effects were also associated with higher levels of anxiety. Given sex differences in effects of risk on BOLD responses, the canonical correlations may relate to sex differences in the expression of psychopathology. For example, differing brain adaptations in males versus females could be responsible for different outcomes in response to stress. Specifically, ADHD is more prevalent in males^68^, whereas anxiety disorders are more prevalent in females^69^. Thus, males and females in this study with greater levels of both heritable and environmental risk demonstrated sex-specific brain adaptations that appear to be advantageous in reducing particular negative outcomes. Importantly, such adaptations do not preclude the possibility that these individuals could have poor outcomes in the future, particularly with regards to alcohol and other substance use. In particular, certain brain adaptations in response to stress are proposed to be developmentally adaptive, despite producing vulnerabilities to poor mental health outcomes later in life^70^.

### Sex differences

Biological sex is an important moderator of risk and expression for various mental disorders, likely due to some combination of sex differences in genetics, hormones, and life experiences^71^. The males and females in this study did not demonstrate baseline differences in mental health and substance use measures but did differ in reported sexual abuse. However, the sexual abuse subscale of the CTQ only accounts for a small proportion of variance in the total CTQ scores due to the generally low scores in both sexes and are thus unlikely to fully account for the observed sex effects during the stop-signal task. Although the mechanisms for sex differences in the current study remain speculative, there are notable sex differences in the brain that could account for the observed findings. Numerous sexual dimorphisms in the healthy brain have been reported^72-75^. In particular, males have greater volume in the anterior cingulate cortex^76^, which is a key region involved inhibitory control and also demonstrated sex differences in the current analysis. Although the developmental distribution of estrogen receptors may account for structural differences in the brain^74,76^, there are also sex differences in brain function, perhaps related to circulating sex steroids^77, 78^. For example, during a stop-signal task, men exhibited increased activation of the anterior cingulate cortex and superior frontal gyrus^34^, regions implicated in the current study, and females generally exhibit poorer behavioral control^38^. CM also differentially affects the neuroendocrine stress response in females compared with males^79,80^, which could account for sex differences in brain function related to CM observed in this study. Sex further modulates FH effects on the brain, such that males with FH exhibit enhanced activation of the left inferior frontal gyrus during response inhibition^37^. Thus, differences between the sexes in the effects of risk factors on brain regions involved in inhibitory control may arise from some combination of sex differences in brain structure and function and sex differences in neuroendocrine and neurodevelopmental responses to risk.

### Limitations

Analyses of task-induced BOLD responses generally provide robust predictors of behavioral and psychiatric variables. Although this sample was relatively large, and analyses examined effects of continuous variables to enhance power of statistical analyses, the results will need to be reproduced in a larger sample. The results of this study likely reflect certain features of the study design and sample, such as the exclusion of psychiatric disorders and the inclusion of 18-19-year-old first-year college students. Thus, this sample likely includes more resilient individuals than a community sample. Finally, continued longitudinal assessments, which are being collected on this sample, will be needed to determine whether effects of risk factors on the brain predict future trajectories of alcohol use and other mental health variables into adulthood.

## Conclusions

Family history of AUD and childhood maltreatment had interacting effects on brain function during a stop-signal task that were dependent upon sex. Sex differences in brain responses to risk factors may account for differential expression of psychopathology between the sexes. Efforts to tailor interventions based on factors such as FH, CM, and/or biological sex may be warranted.

## Data Availability

All data produced in the present study are available upon reasonable request to the authors,

## DATA SHARING

The data that support the findings of this study are available from the corresponding author upon reasonable request.

## ACKNOWLEDGEMENTS

This work was supported by NIH award K01AA026334 (AE) and P60AA011605 (CAB). The authors declare no competing financial interests.

